# Functional connectivity of cognition-related brain networks in adults with fetal alcohol syndrome

**DOI:** 10.1101/2023.05.18.23289319

**Authors:** Benedikt Sundermann, Reinhold Feldmann, Christian Mathys, Johanna M. H. Rau, Stefan Garde, Anna Braje, Josef Weglage, Bettina Pfleiderer

## Abstract

**Background:** Fetal Alcohol Syndrome (FAS) can result in substantial cognitive dysfunction. Many of the cognitive functions affected are subserved by few functional brain networks. Functional connectivity (FC) in these networks can be assessed with resting state functional MRI (rs-fMRI). Alterations of FC have been reported in children and adolescents prenatally exposed to alcohol. However, previous reports varied substantially regarding which exact cognitive networks were affected, their interactions, and the directionalities of FC alterations. Despite persisting deficits, no previous studies have examined FC in older individuals. Purpose of this rs-fMRI study was to assess FC within and between cognition-related networks in young adults with FAS.

**Methods:** Cross-sectional study in patients with FAS (n = 39, age: 20.9 ± 3.4 years) and controls without prenatal alcohol exposure (n = 44, age: 22.2 ± 3.4 years). FC was calculated as correlation between cortical regions in ten cognition-related sub-networks. Subsequent modelling of overall FC was based on two-tailed t-tests comparing FC between FAS and controls. Results were subjected to a hierarchical statistical testing approach, first determining whether there is any alteration of FC in FAS (compared with controls) in the full cognitive connectome, subsequently resolving these findings to the level of either FC within each network or between networks, and finally to individual connections. The overall and network-level tests are based on the Higher Criticism (HC) approach for the detection of rare and week effects in high dimensional data. In an additional exploratory time-resolved FC analysis, potential group differences of dynamic FC states were assessed.

**Results:** Comparing FAS subjects with controls, we observed altered FC of cognition-related brain regions globally, within 7 out of 10 networks, and between networks employing the HC statistic. This was most obvious in the dorsal attention A sub-network, followed by the salience / ventral attention A subnetwork. Findings also spanned subcomponents of the fronto-parietal control and default mode networks. None of the single FC alterations within these networks yielded statistical significance in the final high-resolution analysis. The exploratory time-resolved FC analysis did not show significant group differences in the temporal behavior of FC states.

**Conclusions:** FC in cognition-related brain networks was altered in adults with FAS. Effects were widely distributed across these networks, potentially reflecting the diversity of cognitive deficits in these individuals. Findings were pronounced in attention-related networks in line with attentional deficits previously reported. An additional exploratory time-resolved FC analysis did not reveal altered dynamic FC patterns.

## Introduction

Prenatal alcohol exposure (PAE) can negatively affect a wide range of cognitive functions throughout life. These functions include general intelligence, attention, executive functions (including inhibitory control), learning and memory, language, mathematical abilities, social cognition^1, 2^ as well as impulse control^1, 3, 4^. Deficits associated with PAE frequently persist into adulthood^5, 6^. While the term fetal alcohol spectrum disorders (FASD) generally encompasses a broad range of possible conditions related to PAE, only the full picture of characteristic physical (including growth retardation and facial abnormalities), psychological, and cognitive features is termed Fetal Alcohol Syndrome (FAS)^2, 7, 8^.

Brain activity in individual regions underlying specific cognitive functions can be assessed by functional neuroimaging with a wide range of targeted tests^9^. However, a frequent neuroscientific observation is that many higher cognitive functions, such as those negatively affected in FAS, are subserved by activity in few common sets of brain regions, i.e. functional brain networks. These include task-positive networks with overlapping definitions such as the central executive network, cognitive control network or multiple demands network as well as networks closely interacting with them, such as the salience network and default mode network^10–12^. Spontaneous activity and functional connectivity within and between these networks can be examined by resting state functional magnetic resonance imaging (rs-fMRI)^13, 14^. Static functional connectivity (FC) analysis methods identify correlated activity over a full rs-fMRI data acquisition period^14^. They have recently been supplemented by approaches for assessing dynamic or time-varying FC. Such dynamic FC analyses promise a deeper understanding of dynamic interactions of brain regions within and across these functional networks^15–17^ in health and disease. Dynamic FC measures have been associated with differences in individual attentional performance^18, 19^ and impulsivity^20, 21^. For example, altered FC dynamics have been observed in subjects with attention deficit hyperactivity disorder (ADHD)^22–24^.

Few studies have directly investigated resting-state FC within and between brain networks related to higher cognitive functions in individuals with FAS or prenatal alcohol exposure: Focusing on within-network connectivity, Fan et al. observed reduced FC in a subset of regions within the default mode, salience, ventral attention, dorsal attention, and right fronto-parietal executive control networks in children with FASD compared with non-exposed controls. These networks reflect cognitive functions typically affected in children with FASD. Networks not directly related to cognition were, however, not affected^25^. In another study with children and adolescents with FASD, Little et al. mainly described reductions of FC between core regions of the salience and fronto-parietal control networks and regions from other cognition-related networks rather than within networks^26^. In contrast, Ware et al. found lower within-network but higher between-network FC in attention-related networks in children with FASD^27^. All three studies report relatively high overall similarity of FC in cognition-related networks between exposed individuals and controls, while FC group differences had relatively small effect sizes, contrasting with the distinct clinical deficits in these individuals^25–27^. Further rs-fMRI and methodologically related FC studies in individuals with PAE report evidence of altered overall functional brain organization based on global graph-theoretical measures^28–31^ as well as FC alterations of the default mode network^32^ and within networks less directly related to cognitive control^33, 34^. No previous study has, however, explored dynamic FC in FASD. Beyond that, amid studies describing persisting cognitive deficits into adulthood^5, 6^, FC has not been previously examined in adults prenatally exposed to alcohol.

The main goal of this study was therefore to examine functional connectivity in cognition-related functional brain networks in young adults with FAS and to assess whether these patterns are comparable to alterations previously observed in affected children and adolescents.

The following hypotheses should be tested:

- Static FC in the connectome of all brain regions constituting cognition-related brain networks is altered in young adults with FAS compared with a control group without prenatal alcohol exposure (omnibus test, bi-directional effects possible).
- Static FC within individual cognition-related brain networks is altered in FAS subjects compared with controls without prenatal alcohol exposure (bi-directional effects possible).
- Static FC between cognition-related brain networks is altered in FAS subjects compared with controls without prenatal alcohol exposure (bi-directional effects possible).

In an additional exploratory analysis, we addressed dynamic interactions between cognition-related brain regions in FAS subjects compared with non-exposed controls. This dynamic FC analysis^17^ focused on transitions between putative FC states. Rationale of this analysis is, that less stable FC states in FAS might underlie impaired impulse control (similar to reports in ADHD^22–24^).

## Materials and Methods

This study was approved by the ethics committee of the University of Münster and the Westphalian Chamber of Physicians in Münster. All study procedures were carried out after obtaining written informed consent and in accordance with the Declaration of Helsinki. Samples overlapped with previously published task-based fMRI studies on inhibitory control^35, 36^. The study was carried out in a research setting outside routine clinical care.

### Subjects

Young adult subjects (n = 50) with FAS were initially recruited based on standardized inclusion (including diagnosis of FAS made by a specialist based on the Majewski criteria^37^, 18 to 32 years of age) and exclusion (contraindications for MRI, severe psychiatric, neurological or medical conditions, pregnancy, and severe sensory impairments) criteria. Psychiatric comorbidity or medication in general were not defined as primary exclusion criteria for the FAS group since they are common in individuals with a history of prenatal alcohol exposure^38^. After data acquisition further subjects were excluded after review of potentially biasing medication, structural brain lesions, and MRI data quality control. Two subjects were excluded from the analyses due to use of potentially psychoactive anti-allergic medication unrelated to FAS. In four subjects, no fMRI data were acquired because of claustrophobia. One subject was excluded because of a callosal hypoplasia leading to structural image misregistration. Data from further 4 patients were excluded because of excessive head motion (see section “pre-processing” for criteria). All results are based on the remaining 39 FAS subjects. Current intake of the following potentially psychoactive medication was reported in the FAS group: methylphenidate or derivatives (n = 8), antipsychotics (n =5), and antidepressants (n = 1).

The control group consisted of subjects without a history of prenatal alcohol exposure. Initially, n = 52 subjects were recruited. Apart from general exclusion of subjects with psychoactive medication in the control group, inclusion and exclusion criteria (both for initial inclusion and after data acquisition) were identical in both groups. Two subjects were excluded due to use of psychoactive medication. One subject was excluded because of a large frontal venous anomaly^39^. One fMRI dataset was excluded because of a technical failure. Data from 4 subjects were excluded because of excessive head motion. We observed a statistically significant age difference between the groups with a small effect size (Table 1). We refrained from excluding further control subjects in order not to compromise statistical power considering that only small age effects on FC are expected in this particular age range^40^. Finally, 44 controls were included in further analyses. Demographical data of the final sample are reported in Table 1.

**Table 1.** Demographical and clinical characteristics of participants with FAS and controls.

| a) |  | Total | Female | Male | p<sup>a</sup> |
| --- | --- | --- | --- | --- | --- |
| Sex | FAS | 39 | 17 (43.6 %) | 22 (56.4 %) | .319 |
|  | CON | 44 | 24 (54.5 %) | 20 (45.5 %) |  |
| b) | Group | Mean ± SD | Median | Range | p<sup>b</sup> |
| Age (years) | FAS | 20.9 ± 3.4 | 20 | 18 - 32 | .013* |
|  | CON | 22.2 ± 3.4 | 21 | 18 - 32 |  |
| EHI<br>Handedness<br>index | FAS | 71.6 ± 37.9 | 83.3 | -58.3 - 100 | .887 |
|  | CON | 69.8 ± 40.3 | 83.3 | -58.3 - 100 |  |
| TMT<br>reaction<br>time (sec) | FAS | 97.2 ± 36.7 | 86.3 | 54.3 - 220.0 | < .001* |
|  | CON | 54.0 ± 8.7 | 54.1 | 35.5 - 74.8 |  |
| IQ (mean ±<br>SD)<sup>c</sup> | FAS<sup>d</sup> | 82.0 ± 16.7 | 80.5 | 58 - 116 | < .001* |
|  | CON | 117.7 ± 15.0 | 116.5 | 88 - 150 |  |

<sup>a</sup> Chi-squared test (sex distribution between groups), <sup>b</sup> Mann-Whitney-U-test, <sup>c</sup> based on TMT reaction times, <sup>d</sup> only calculated for n = 36 within valid range, \* significant difference. FAS: fetal alcohol syndrome, CON: controls, SD: standard deviation, NA: not applicable, EHI: Edinburgh Handedness Inventory, TMT: trail-making task, IQ: intelligence quotient estimated from TMT performance

Statistical tests on clinical and demographical data were carried out in SPSS (version 27.0, IBM, Armonk, NY, USA).

### Neuropsychological pre-assessment

Subjects completed questionnaire-based pre-tests for handedness (Edinburgh Handedness Inventory, EHI)^41^, processing speed (trail-making task, TMT)^42^, and screening for severe mental comorbidity (DIA-X Stamm-Screening questionnaire, SSQ)^43^. A general intelligence estimate was calculated based on TMT results^42, 44^. Quantitative test results are presented in Table 1.

### Acquisition of MRI data

MRI data were acquired at 3 Tesla (Intera with Achieva upgrade, Philips, Best, NL). FMRI data were acquired during 9:45 min of wakeful rest using gradient echo planar imaging covering the whole brain (234 functional volumes after 5 non-recorded dummy scans to allow for signal equilibration; repetition time: 2500 ms, echo time: 35 ms, 36 axial slices, spatial resolution 3.6 x 3.6 x 3.6 mm). T1-weighted 3D data were acquired with an inversion-prepared turbo field echo (TFE) sequence (inversion time: 411 ms, repetition time: 7.1 ms, echo time: 3.5 ms, flip angle: 9°, sagittal slices measured with 2 mm thickness, reconstructed spatial resolution by zero-filling in k-space 1.0 x 1.0 x 1.0 mm)

### Analysis of MRI data

#### Pre-processing and image quality control

MRI data were converted to the Brain Imaging Data Structure (BIDS)^45^ using in-house scripts preceding the BiDirect-BIDS-ConverteR^46^. Facial features were removed from the T1-weighted anatomical data^47^. Main MRI data pre-processing was carried out using fMRIPrep^48^ (version 20.0.7) briefly consisting of motion estimation and correction, co-registration of fMRI and structural MRI data, estimation of noise regressors as well as standard space normalisation. Please consult the Supplementary Methods for further details. Subsequent actual denoising was carried out using fMRIDenoise^49^ (version 0.2.1), comprising regressing out 24 head motion parameters (3 translations, 3 rotations, their 6 temporal derivatives, and their 12 quadratic terms)^50^, 8 physiological noise parameters (mean physiological signals from white matter and cerebrospinal fluid, their 2 temporal derivatives, and 4 quadratic terms)^50^ as well as movement spike regression based on frame-wise displacement (FD > 0.5 mm) and so-called “DVARS” (> 3) thresholds^51^, temporally filtering (0.008 – 0.08 Hz), and, finally, smoothing the resulting standard space image with a Gaussian kernel (FWHM = 6 mm). This pre-processing leads to denoised fMRI data in a common standard space as input for further analyses.

The following steps were taken for MRI data quality control (numbers of excluded participants reported in the section “subjects”): Structural MRI data were screened by a radiologist for incidental findings and major artefacts. fMRIPrep reports were reviewed for registration errors and image artefacts. Subject exclusion for excessive head motion (see section “Subjects”) was based on pre- processing criteria (mean FD > 0.3 mm or maximum FD > 5 mm or more than 20% outlier data points). FD did not differ significantly between FAS and controls. However, there was a trend towards higher mean and maximum FD in the FAS group (see Supplementary Table 1).

#### Static functional connectivity analysis: general approach

Functional connectivity analyses were based on a cortical atlas (“Schaefer atlas”) derived from rs-fMRI data in 1489 subjects. The atlas was obtained from TemplateFlow to match the dimensions of the fmriprep outputs^52^. The atlas version with 400 parcels adopted here is the most extensively validated version of this atlas, e.g. regarding stability and correspondence with markers of brain function^53^. The individual parcels in the published atlas have been matched to 17 non-overlapping networks from a previously established atlas by Yeo et al.^54^. 10 cognition-related components out of these 17 networks were selected for further analysis: the dorsal attention network (2 sub-networks A and B), the salience / ventral attention network (2 sub-networks A and B), the mainly fronto-parietal control network (3 sub-networks A-C), and the default-mode network (3 sub-networks A-C). Timeseries extraction from the pre-processed fMRI data and calculation of z-transformed Pearson correlation coefficients as primary measures of FC were carried out with the Data Processing Assistant for Resting-State fMRI (DPARSF, version 5.2)^55^ based on Matlab 2019b (The MathWorks, Natick, MA, USA). As a basis for subsequent modelling, we carried out multiple two-tailed two-sample t-tests (one test per pair of regions), comparing z-transformed correlation coefficients among regions of interest between FAS patients and controls.

We followed a hierarchical statistical testing approach with three levels of analysis: 1) first, determining whether there is any alteration of FC in FAS subjects compared with controls in the full connectome of 243 regions constituting these 10 cognition-related networks (i.e. an omnibus test) globally and subsequently aiming to resolve these findings, 2) to the level of either FC within each network or between-network connectivity, and finally 3) to individual connections.

#### Static functional connectivity analysis: global analysis

The omnibus test on the full connectome is based on the “Higher Criticism” (HC) approach^56^ in an improved version^57^ implemented in Matlab (https://www.stat.cmu.edu/~jiashun/Research/software/HC/). HC statistics can be applied in order to test whether there are any non-zero effects within a large number of individual tests carried out in high-dimensional datasets. They are thus suitable to identify the existence of rare and weak effects in such data^56^. HC follows the rationale of p-value histogram analyses: Under a null-hypothesis of only zero-effects in multiple parallel tests, an equal distribution of p-values is expected. Under the alternative hypothesis of existing non-zero effects, there is an excess of low p-values^58^. In simplified terms, HC statistics test a joint hypothesis of such an excess of low p-values^56, 58^. HC has been increasingly popular for detecting effects in high-dimensional data such as in genetic^56^ and economic^59^ research. It has been argued that HC could be favorable for the detection of rare events compared with conventional false-discovery rate (FDR) or family-wise error rate (FWE) correction methods^56^. Considering the similarly high dimensionality of FC datasets, HC has recently been applied to rs-fMRI analyses^60^.

#### Static functional connectivity analysis: within-network HC analysis

Subsequently, we aimed to determine which of the 10 cognition-related networks were affected by within-network functional connectivity differences between FAS and control subjects. We therefore carried out equivalent HC tests separately for these networks. Each set of tests included the full set of correlation coefficients between all regions within each network.

#### Static functional connectivity analysis: between-network HC analysis

For a similar analysis of between-network FC (i.e., to determine whether any between-network FC differences were present), we concatenated all parcels for each of the 10 networks separately. This resulted in a single mask for each network before time-series data extraction. The resulting correlation coefficients were assessed with an equivalent HC test.

#### Static functional connectivity analysis: analysis of individual connections

In a third level we aimed to identify single between- and within-network connections exhibiting statistically significant FC differences between FAS subjects and controls. Therefore, in contrast to the previously described HC-based joint-hypothesis tests, we now FDR-adjusted^61^ the individual hypothesis tests of between- and within-network connectivity (q < 0.05), using an FDR implementation in Matlab (https://brainder.org/2011/09/05/fdr-corrected-fdr-adjusted-p-values). This was carried out separately for either all 45 between network connections or all individual connections within each network.

#### Exploratory time-resolved functional connectivity analysis

Beyond the static FC analysis, we carried out an exploratory dynamic FC analysis, using a sliding window approach with the DynamicBC toolbox (version 2.2)^62^. FC between all 243 regions in the cognition-related networks was calculated for each individual subject by Pearson linear correlation separately in overlapping windows with a length of 18 consecutive functional volumes equivalent to 45 s, similar to window lengths in previous studies^63, 64^, and with a 60 % overlap. The resulting time-resolved FC estimates from individual time windows were grouped by similarity (K-means cluster analysis, distance measure: correlation) in order to derive presumed FC states in the entire sample. Established methods were used to estimate the optimal number of clusters and assess the goodness of fit of the clustering solutions: (1) The optimal number of clusters (search range: 2 to 10) was estimated using the Calinski-Harabasz^65^ and Davies–Bouldin^66^ indices resulting in 2 clusters (see Supplementary Fig. 2). (2) Cluster-separability was estimated by a silhouette analysis. Briefly, the silhouette analysis assesses, how similar an individual element is to other elements within its own cluster compared with elements in other clusters^67^. The following summary measures describing the temporal dynamics of putative FC states were calculated for individual subjects: number of transitions (NT) between connectivity states, mean dwell time (MDT) per cluster, and frequency of observing each cluster (FRC). Subjects with FAS and controls were compared regarding NT and MDT using t-tests or Mann-Whitney U tests.

## Results

### Static functional connectivity analysis: global analysis

We observed significantly altered FC of cognition-related brain regions in FAS subjects compared with non-exposed controls in the global (all parcels of all 10 cognition-related networks) analysis over the entire data acquisition period. This finding is based on a joint hypothesis test (HC test statistic: 39.02) which aims to detect the existence of alterations within this high dimensional dataset but which does not identify which exact connections are altered (Fig. 1). Quantitative results of FC group differences in connections between single atlas regions as depicted in Fig. 1C are shared (not currently available, to be added as supplementary material or in a repository at the time of publication depending on journal policy).

**Fig. 1.**
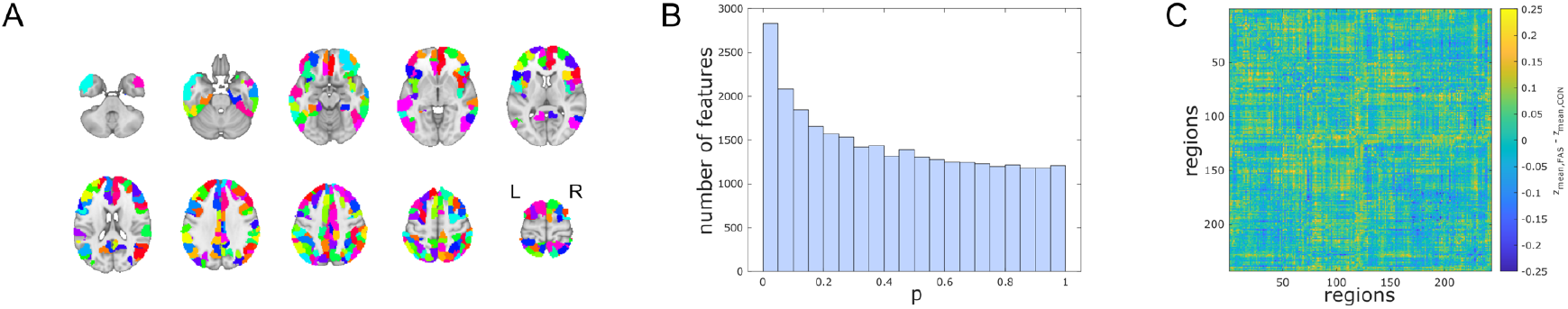
Global analysis of static functional connectivity of cognition-related brain networks. A) Full atlas-based selection of 243 individual brain regions in cognition-related networks (redundant color coding for illustration of atlas resolution only). B) P-value histogram of multiple individual two-sided t-tests comparing functional connectivity among all these brain regions between FAS patients and control subjects. Under the null-hypothesis of equal functional connectivity in both groups, equal numbers of p-values are expected in each histogram bin. The histogram shows an excess of low p-values. The existence of at least rare and/or weak effects is also confirmed by a test of the joint hypothesis based on higher criticism statistics. This means that regarding a significant number of functional connections, FAS patients differ from healthy controls. C) Unthresholded matrix of connectivity group differences describing the full connectome of cognition-related brain regions. Yellow: mean z-transformed correlation coefficients relatively increased in FAS compared with controls. Blue: relatively decreased functional connectivity in FAS.

### Static functional connectivity analysis: within-network HC analysis

FC was altered within 7 out of 10 of these cognition-related brain networks based on the joint hypothesis tests. Based on the HC test statistic and supported by p-value histograms of tests of individual functional connections, this effect was most obvious in the dorsal attention A sub-network (HC test statistic: 12.33), followed by the salience / ventral attention A network (HC test statistic: 9.17). However, findings also spanned sub-networks B and C of the fronto-parietal control (HC test statistics: 5.33 and 4.29) and sub-networks A, B, and C of the default mode (HC test statistics: 3.80, 6.42 and 4.33) networks (Fig. 2).

**Fig. 2.**
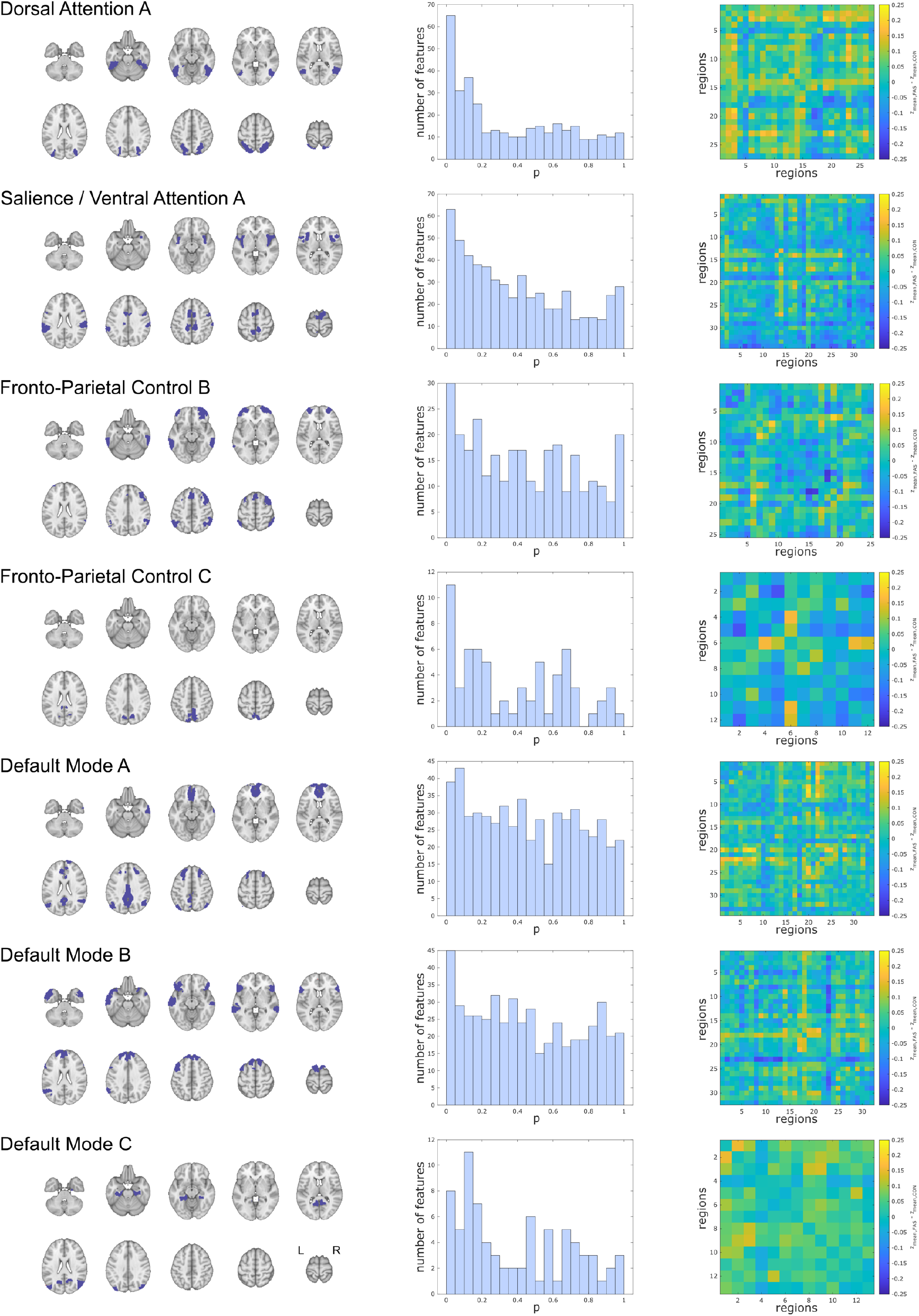
Within-network static functional connectivity of cognition-related brain networks. Seven (out of ten) sub-networks exhibiting altered functional connectivity in FAS patients compared with controls. Left column: Overview of the networks’ overall extent. Middle column: P-value histograms (different scaling reflecting different numbers of regions in each network) of multiple two-sided t-tests comparing functional connectivity within these sub-networks between FAS patients and control subjects. Under the null-hypothesis of equal functional connectivity in both groups, equal numbers of p-values are expected in each histogram bin. The histogram shows an excess of low p-values, confirmed by a test of the joint hypothesis based on higher criticism statistics. This means that FAS patients differ from healthy controls regarding at least rare and/or weak effects. Right column: Unthresholded matrices of connectivity group differences describing the full connections of cognition- related brain regions within each network. Yellow: mean z-transformed correlation coefficients relatively increased in FAS compared with controls. Blue: relatively decreased functional connectivity in FAS. The remaining three networks without significant results are presented in Supplementary Fig. 1.

### Static functional connectivity analysis: between-network HC analysis

A subsequent analysis revealed altered FC between cognition-related brain networks based on an equivalent joint hypothesis test (HC test statistic: 4.50). Descriptively, underlying strongest relative decreases of FC (ranking of correlation coefficient group differences) in FAS subjects were observed between the salience / ventral attention B and fronto-parietal control C sub-networks. The strongest relative increases were observed between the dorsal attention B and fronto-parietal control sub- networks as well as between the default mode C sub-network and other parts of the default mode network and fronto-parietal control network (Fig. 3).

**Fig. 3.**
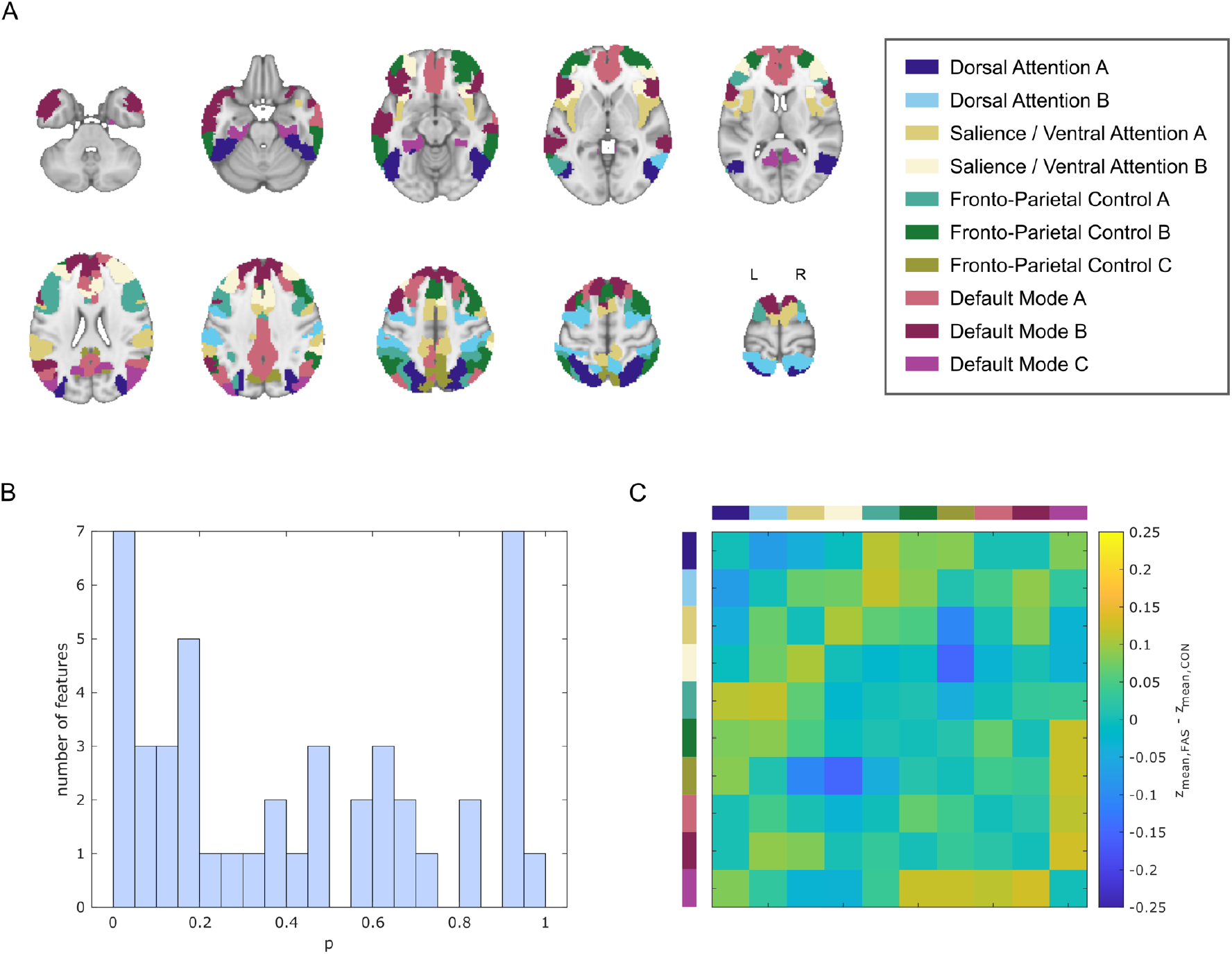
Between-network static functional connectivity of cognition-related brain networks. **A)** 10 cognition-related networks. Each color represents an individual sub-network (network-wise concatenation of individual regions based on atlas labels). B) P-value histogram of multiple two-sided t-tests comparing functional connectivity among these sub-networks between FAS patients and control subjects. Under the null-hypothesis of equal functional connectivity in both groups, equal numbers of p-values are expected in each histogram bin. The histogram shows an excess of low p-values confirmed by a test of the joint hypothesis based on higher criticism statistics. This means that regarding a significant number of functional connections, FAS patients differ from healthy controls. C) Unthresholded matrix of connectivity group differences describing the full connectome of cognition- related brain regions. Yellow: mean z-transformed correlation coefficients relatively increased in FAS compared with controls. Blue: relatively decreased functional connectivity in FAS.

### Static functional connectivity analysis: analysis of individual connections

None of the individual FC alterations (either from the within- or between-network analysis) were statistically significant when correcting the separate tests of individual connections for multiple comparisons.

### Exploratory time-resolved functional connectivity analysis

In the exploratory dynamic FC analysis, a solution consisting of two FC states was empirically derived as the optimal number of clusters across the entire sample. These two clusters representing putative FC states were only weakly separable (see silhouette values and further goodness-of-fit statistics for the clustering solutions in Supplementary Fig. 2). Both clusters differed mainly regarding (1) the relatively connectivity strength of the DMN and (2) the extent that the DMN appeared interconnected with other cognition-related brain networks. See Supplementary Fig. 3 for further details on the clusters. FAS subjects did not differ significantly regarding the temporal dynamics of these FC states NT, MDT, and FRC (Table 2).

**Table 2.** Group comparison results for the optimal clustering solution (2cl) of the time-resolved functional connectivity analysis. Clusters represent estimates of putative FC states. In summary, cluster 1 represents widely distributed FC dominated by the DMN while cluster 2 exhibits stronger dichotomisation between the DMN and the other cognition-related networks.

|  | FAS | CON | p |
| --- | --- | --- | --- |
| Number of transitions<sup>a</sup> | 6.59 ± 2.99 | 7.11 ± 2.24 | 0.366 |
| Mean dwell time (cluster 1)<sup>b</sup> | 3.33 (0.00-13.5) | 3.29 (1.33-27.00) | 0.809 |
| Mean dwell time (cluster 2)<sup>b</sup> | 3.00 (1.00-28.00) | 3.00 (1.00-9.00) | 0.982 |
| Frequency (cluster 1)<sup>c</sup> | 57.14 (0-96) % | 54.57 (11-96) % | 0.780 |
| Frequency (cluster 2)<sup>c</sup> | 42.86 (4-100) % | 46.43 (4-89) % | 0.780 |

<sup>a</sup> mean ± standard deviation and t-test result), <sup>b</sup> unit: number of (partially overlapping) windows, median (range) and Mann-Whitney-U-test result, <sup>c</sup> median (range) and Mann-Whitney-U-test result. FAS: fetal alcohol syndrome, CON: controls

## Discussion

In summary, FC was altered in adults with FAS compared to controls not exposed to alcohol prenatally both within a majority of cognition-related networks (including the dorsal attention network and, to a lesser degree, the salience / ventral attention networks, the fronto-parietal control network, and the default mode network) and between these networks. FC changes were widely distributed across networks. These results on the global and network level are based on an HC approach, indicating that at least rare and weak group effects seem to be present within and between these networks^56^. HC-based findings do, however, not necessarily mean that FC is changed in a majority of connections within and between these networks. Group effects could not be further resolved to connections between individual regions using conventional mass-univariate testing with multiple comparison correction. In the additional exploratory time-resolved FC analysis, altered FC dynamics in the FAS group could not be observed.

The wide distribution of findings across cognition-related networks is in line with the similarly wide range of cognitive deficits observed in individuals with FAS^1^. It thus suggests a rather distributed neural basis (i.e. unspecific alcohol-related damage) of such deficits rather than strongly localized alterations. A similarly wide distribution of FC alterations across networks was observed by Fan et al. in children with FASD based on an independent component analysis with dual regression^25^. Thus, this observation of a wide distribution spans different age groups (from childhood to young adulthood) and rs-fMRI analysis approaches. Visual interpretation of the connectivity matrices reveals different directionalities of findings: Some connections exhibited higher, and some lower FC in FAS. This bidirectionality is generally in line with a study in children with FASD by Ware et al.; They also report different directionalities of FC alterations^27^. Consequently, a simple picture of either overall increases or decreases of FC in FAS does not exist.

The observation of more obvious effects in attention- and salience-related systems compared with networks underlying other cognitive functions highlights the importance of attentional deficits in FASD^1, 68–72^, including adults^5, 73^. However, this interpretation might be somewhat limited by the infeasibility of a direct quantitative comparison between networks as well as reverse inference^74, 75^. Our rs-fMRI findings are generally in line with previous studies suggesting a particular involvement of attentional functions and underlying neural systems in FAS when compared to other cognitive deficits: Response and activation patterns in a Go/NoGo task in a sample of young female adults overlapping with this study also provide indirect evidence of a particular importance of attentional deficits compared with inhibitory control deficits in this age group^36^. Attention-related networks were also altered in other alcohol-exposed samples studied with fMRI: Attention networks were among those altered in studies by Fan et.al^25^ and Ware et al^27^. In the latter study, FC alterations were associated with differences in attentional performance measures. The authors consequently conclude that the patterns observed (lower within-network, higher between-network FC) provide support for reduced attention network specialization and inefficiency^27^. Reduced FC between key regions of the salience network with other cognition-related networks were among the key findings by Little et al.^26^. A resting-state magnetoencephalography study provided initial evidence that spectral changes in subjects with a history of prenatal alcohol exposure might be related to a subset of attention-related measures but also to emotional behavior^76^. Diffusion tensor imaging revealed reduced inter-network structural connectivity including the ventral attention and default mode network in children and adolescents prenatally exposed to alcohol^77^.

To a lesser extent, we also observed altered FC in parts of the fronto-parietal control network. This network is considered a flexible hub that interacts with other processing networks in order to orchestrate performance in a wide range of cognitive tasks^78^. Changes in fronto-parietal network FC were among those also observed in younger subjects with FASD^25, 26^. There is further evidence of altered activity in these networks in children with FASD from task-based fMRI studies on inhibitory control^79^ and working memory^80^.

The default mode network (DMN), though classically reported as anti-correlated with task-positive cognitive networks^81^, is considered to be involved in cognitive functions including task-switching and integration of information^82, 83^. There is evidence of regional differentiation within the DMN, with subdivisions subserving different cognitive functions^84^. Fan et al observed altered FC in the anterior part of the DMN, discussed as subserving social perception, judgment, and self-referential processing in individuals with FASD; however, they found no changes within the posterior DMN^25^. Correspondingly, we predominantly observed FC alterations affecting the “DMN B” sub-network of the DMN, which also mainly includes anterior subregions of the DMN. Beyond that, there is further evidence of less regionally specific DMN dysfunction in FASD from a seed-based FC analysis^32^.

The main static FC analysis applied in this study follows a hierarchical statistical approach, partially based on HC statistics. This approach addresses the general limitations of functional neuroimaging analyses in relatively infrequent disorders such as FAS (estimated global prevalence of FAS: 14.6 per 10 000 people^85^): Conventional high-dimensional FC analysis methods, such as frequently-used mass-univariate statistical testing, carry the risk to report only a “tip of the iceberg” of true underlying alterations due to lower than optimal statistical power. There is an increased risk of both published findings being false-positive^86, 87^ or false-negative findings^86^. Further in-depth discussions on this issue have been published^87–89^. Consequently, statistical thresholding of mass-univariate analyses (mainly for multiple comparison correction) might in part explain ambiguous FC results in previous studies in children with FASD^25–27^. There are increasing efforts to report subthreshold effects in fMRI studies in order to facilitate better interpretation of underlying patterns^90–92^. An example is the additional presentation of unthresholded activation or connectivity maps^91, 93^. Our hierarchical approach with HC-based joint hypothesis tests^56, 57^ at the network level might help avoid these shortcomings without sacrificing information from individual connections. Compared with conventional mass-univariate fMRI analyses, it avoids selectively reporting and interpreting few selected results that would pass a multiple-comparison threshold but might not well represent the true underlying effect in a medium-power setting. Please see the “methods” section for a more detailed description of the approach and underlying rationale, as well as the “limitations” section for further methodical aspects.

Our findings discussed so far are based on conventional static FC analyses. Clinical features typically observed in individuals prenatally exposed to alcohol include impulsivity or hyperactivity (including overlap/comorbidity with attention deficit hyperactivity disorder)^1, 3, 4^. These clinical features might suggest a potential dynamic or temporally-changing nature of underlying neural disease mechanisms. This assumption is supported by dynamic FC alterations previously reported in ADHD^22–24^. Contrary to this assumption, we did not observe alterations of features representing non-stationarity in the exploratory time-resolved analysis of dynamical aspects of FC. In particular, compared to controls FAS subjects did neither change more or less frequently between two putative FC states, nor did they remain in the different FC states for shorter or longer periods of times. Thus, we did not find evidence of altered dynamic interactions of brain regions of different networks. There is at least some evidence in children, that hyperactivity might be less severe in PAE compared with ADHD^94^. Though those findings cannot be directly translated to adult FAS subjects, relatively lower hyperactivity might be the reason for the lack of dynamic FC alterations observed in our study. These findings are also generally in line with a previous rs-fMRI study reporting no alteration in regional temporal variance in children with low levels of PAE, although this study differs substantially from our dynamic FC analysis regarding disease severity, age and analysis method^95^.

### Potential limitations

Findings of this first FC analysis in adults with PAE are restricted to young adults (ages 18-32 years) with FAS. They do not necessarily translate to other age groups. Results of this HC analyses aiming at detecting rare and weak effects might, in part, be influenced by subliminal demographic effects, despite the relatively homogeneous sample in a narrow age range. Furthermore, findings do also not necessarily translate to other gradations across the FASD spectrum. Despite greater psychopathology, attention deficits, and impulsiveness compared with controls, a recent study did not find network-based FC alterations in a population of adolescents with a wide range of PAE, i.e. less severely exposed individuals^96^.

Although precautions were taken to minimize head motion and even though measures did not differ significantly between groups, it cannot be fully excluded that parts of the results are movement-related. We have adopted state-of-the-art motion-correction methods during data preprocessing (see the ‘Methods’ section) to further avoid a potential movement bias.

Statistical power of the final step of the hierarchical analysis approach (resolving FC alterations to single connections between pairs of regions) is potentially limited by the high dimensionality of the underlying atlas. This atlas resolution was chosen because it is extensively validated and aims to approach neurobiological estimates of the number of truly distinct cortical regions^53^. FC analyses based on atlases with higher dimensionality similar to this have been shown to better reflect the brain’s functional architecture compared with traditional atlases based on surface anatomy^97^. The HC-based approach was chosen here instead of e.g. region-of-interest concatenation (averaging) to allow global and network-wise inference while maintaining the advantages of the high resolution atlas. Findings are limited to the cortex and do not include subcortical gray matter nuclei within these networks^98, 99^. The functional network labels provided by the Schaefer atlas^53^ are a prerequisite for this approach. Additionally, it has to be noted that there is an ongoing debate about functional network nomenclature, so that the networks described here may deviate from studies using other brain atlases^100^.

Though widely used in other research areas with high dimensional data^56, 59^, the HC statistic has only recently been introduced to fMRI^60, 92^. The HC statistic primarily assumes statistically independent features, since correlations among features can lead to unbalanced p-value histograms, however without expecting peaks in the first histogram bin (low p-values). It has thus been argued that the influence of correlations among features is negligible when the underlying histograms show typical behavior^58^. In addition to the HC statistic, we therefore visually interpreted the underlying p-value histograms as a plausibility control and observed well-behaved p-value histograms in the global and within-network analysis and to a slightly lesser degree in the between-network analysis. The HC-based global hypothesis test is not part of the statistical framework resulting in p-values for determining statistical significance. Thus, no exact p-values can be provided. However, p-values for typical HC values have been approximated, e.g. p = 0.05 for an HC threshold of 4.83 and p = 0.01 for an HC threshold of 10.0 ^101^. Even when using these approximated p-values, thus departing from the notion of the original HC statistic, a majority of results would be statistically significant at the p < 0.05 level or stricter p-values.

Dynamic or time-resolved FC analysis is a promising, already widely used, yet still evolving rs-fMRI analysis method^15–17^. Thus, there is currently a relatively high methodological variability in this field^15, 17^. Here, we adopted a widely used sliding-window approach^17^ and refrained from extensively exploring analysis settings in order to avoid false positive findings^102^. Hence, there is a risk to miss group differences of FC dynamics which might have been uncovered with other, less well-established dynamic FC analysis approaches which might address limitations of the sliding window approach^15, 17, 103^. Similar to methodological heterogeneity there is still no consensus of connectivity states to be expected in a normal population. However, a relatively small number of FC states partially reflecting changing interactions of parts of the DMN, similar to those observed here (Supplementary Fig. 3) has been repeatedly reported^17, 104^. Silhouette values (Supplementary Fig. 2) indicate that clusters observed in our analysis may not be well separated. Thus, these two clusters capture dynamic FC changes as a model but might not represent truly discrete FC states in a neurobiological sense. Cluster frequencies suggest a high inter-subject variability. Despite these general limitations of this evolving methodology, we believe that our exploratory approach can be a starting point for further investigations on dynamic FC in FAS and other disorders with impaired brain development.

## Conclusions

We observed altered FC in cognition-related brain networks in young adults with FAS. Using a HC-based statistical approach, this study provides evidence of the existence of at least rare and weak effects (i.e, FC differences between subjects with FAS and controls) widely distributed across a majority of these networks, potentially underlying the diversity of cognitive deficits in these individuals. Findings were pronounced in attention-related sub-networks, which is in line with substantial attentional deficits previously reported. An exploratory time-resolved analysis, however, did not identify altered FC dynamics and could thus not explain reduced impulse control and attention deficits which have been frequently reported in FAS.

## Data Availability

On request to the authors, further intermediate data on a level independent from the individual subjects can be shared. Due to German data protection regulations and to safeguard subject confidentiality, data on the level of individual subjects cannot be made available. 

## Data availability

On request to the authors, further intermediate data on a level independent from the individual subjects can be shared. Due to German data protection regulations and to safeguard subject confidentiality, data on the level of individual subjects cannot be made available (no participant consent for sharing these primary data).

## Acknowledgements

We thank all participants for taking part in this study. We acknowledge the kind support of staff at the Translational Research Imaging Center (University Hospital Muenster, Clinic of Radiology). We thank Mahboobeh Dehghan Nayyeri and Niklas Wulms for assistance with BIDS data handling, and Anke McLeod for insightful discussions on the manuscript. The color scheme representing networks in Fig. 3 has been inspired by Paul Tol’s guide on accessible color schemes (https://personal.sron.nl/~pault/data/colourschemes.pdf). Preliminary results of this study have been accepted for presentation at the 2023 annual meeting of the International Society for Magnetic Resonance in Medicine (ISMRM), program number 3336.

## Supplementary material

Supplementary material included at the end of the preprint PDF.

## Supplementary material

### Supplementary methods

#### MRI data preprocessing

The following fMRIPrep “boilerplate” (indented text) describes the preprocessing steps in detail. The text has intentionally been left completely unchanged according to the fMRIPrep recommendations for optimal reproducibility. Please note that fMRIPrep generated multiple preprocessing outputs which could be used in different denoising and analysis strategies. Not all of these parallel outputs have been used for further processing in this study. Details about which outputs were used for actual denoising and further functional connectivity modelling are presented in the main text.

Results included in this manuscript come from preprocessing performed using *fMRIPrep* 20.0.7 (Esteban, Markiewicz, et al. (2018); Esteban, Blair, et al. (2018); RRID:SCR_016216), which is based on *Nipype* 1.4.2 (Gorgolewski et al. (2011); Gorgolewski et al. (2018); RRID:SCR_002502).

#### Anatomical data preprocessing

The T1-weighted (T1w) image was corrected for intensity non-uniformity (INU) with N4BiasFieldCorrection (Tustison et al. 2010), distributed with ANTs 2.2.0 (Avants et al. 2008, RRID:SCR_004757), and used as T1w-reference throughout the workflow. The T1w-reference was then skull-stripped with a *Nipype* implementation of the antsBrainExtraction.sh workflow (from ANTs), using OASIS30ANTs as target template. Brain tissue segmentation of cerebrospinal fluid (CSF), white-matter (WM) and gray-matter (GM) was performed on the brain-extracted T1w using fast (FSL 5.0.9, RRID:SCR_002823, Zhang, Brady, and Smith 2001). Brain surfaces were reconstructed using recon-all (FreeSurfer 6.0.1, RRID:SCR_001847, Dale, Fischl, and Sereno 1999), and the brain mask estimated previously was refined with a custom variation of the method to reconcile ANTs-derived and FreeSurfer-derived segmentations of the cortical gray-matter of Mindboggle (RRID:SCR_002438, Klein et al. 2017). Volume-based spatial normalization to one standard space (MNI152NLin2009cAsym) was performed through nonlinear registration with antsRegistration (ANTs 2.2.0), using brain-extracted versions of both T1w reference and the T1w template. The following template was selected for spatial normalization: *ICBM 152 Nonlinear Asymmetrical template version 2009c* [Fonov et al. (2009), RRID:SCR_008796; TemplateFlow ID: MNI152NLin2009cAsym],

#### Functional data preprocessing

For each of the 1 BOLD runs found per subject (across all tasks and sessions), the following preprocessing was performed. First, a reference volume and its skull-stripped version were generated using a custom methodology of *fMRIPrep*. Susceptibility distortion correction (SDC) was omitted. The BOLD reference was then co-registered to the T1w reference using bbregister (FreeSurfer) which implements boundary-based registration (Greve and Fischl 2009). Co- registration was configured with six degrees of freedom. Head-motion parameters with respect to the BOLD reference (transformation matrices, and six corresponding rotation and translation parameters) are estimated before any spatiotemporal filtering using mcflirt (FSL 5.0.9, Jenkinson et al. 2002). The BOLD time-series (including slice-timing correction when applied) were resampled onto their original, native space by applying the transforms to correct for head-motion. These resampled BOLD time-series will be referred to as *preprocessed BOLD in original space*, or just *preprocessed BOLD*. The BOLD time-series were resampled into standard space, generating a *preprocessed BOLD run in MNI152NLin2009cAsym space*. First, a reference volume and its skull-stripped version were generated using a custom methodology of *fMRIPrep*. Several confounding time-series were calculated based on the *preprocessed BOLD*: framewise displacement (FD), DVARS and three region-wise global signals. FD and DVARS are calculated for each functional run, both using their implementations in *Nipype* (following the definitions by Power et al. 2014). The three global signals are extracted within the CSF, the WM, and the whole-brain masks. Additionally, a set of physiological regressors were extracted to allow for component-based noise correction (*CompCor*, Behzadi et al. 2007). Principal components are estimated after high-pass filtering the *preprocessed BOLD* time-series (using a discrete cosine filter with 128s cut-off) for the two *CompCor* variants: temporal (tCompCor) and anatomical (aCompCor). tCompCor components are then calculated from the top 5% variable voxels within a mask covering the subcortical regions. This subcortical mask is obtained by heavily eroding the brain mask, which ensures it does not include cortical GM regions. For aCompCor, components are calculated within the intersection of the aforementioned mask and the union of CSF and WM masks calculated in T1w space, after their projection to the native space of each functional run (using the inverse BOLD-to- T1w transformation). Components are also calculated separately within the WM and CSF masks.

For each CompCor decomposition, the *k* components with the largest singular values are retained, such that the retained components’ time series are sufficient to explain 50 percent of variance across the nuisance mask (CSF, WM, combined, or temporal). The remaining components are dropped from consideration. The head-motion estimates calculated in the correction step were also placed within the corresponding confounds file. The confound time series derived from head motion estimates and global signals were expanded with the inclusion of temporal derivatives and quadratic terms for each (Satterthwaite et al. 2013). Frames that exceeded a threshold of 0.5 mm FD or 1.5 standardised DVARS were annotated as motion outliers. All resamplings can be performed with *a single interpolation step* by composing all the pertinent transformations (i.e. head-motion transform matrices, susceptibility distortion correction when available, and co-registrations to anatomical and output spaces). Gridded (volumetric) resamplings were performed using antsApplyTransforms (ANTs), configured with Lanczos interpolation to minimize the smoothing effects of other kernels (Lanczos 1964). Non-gridded (surface) resamplings were performed using mri_vol2surf (FreeSurfer).

Many internal operations of *fMRIPrep* use *Nilearn* 0.6.2 (Abraham et al. 2014, RRID:SCR_001362), mostly within the functional processing workflow. For more details of the pipeline, see <u>the section corresponding to workflows in</u> *fMRIPrep*<u>’s documentation</u>.

#### Copyright Waiver

The above boilerplate text was automatically generated by fMRIPrep with the express intention that users should copy and paste this text into their manuscripts *unchanged*. It is released under the <u>CC0</u> license.

## Supplementary tables

**Supplementary Table 1.**
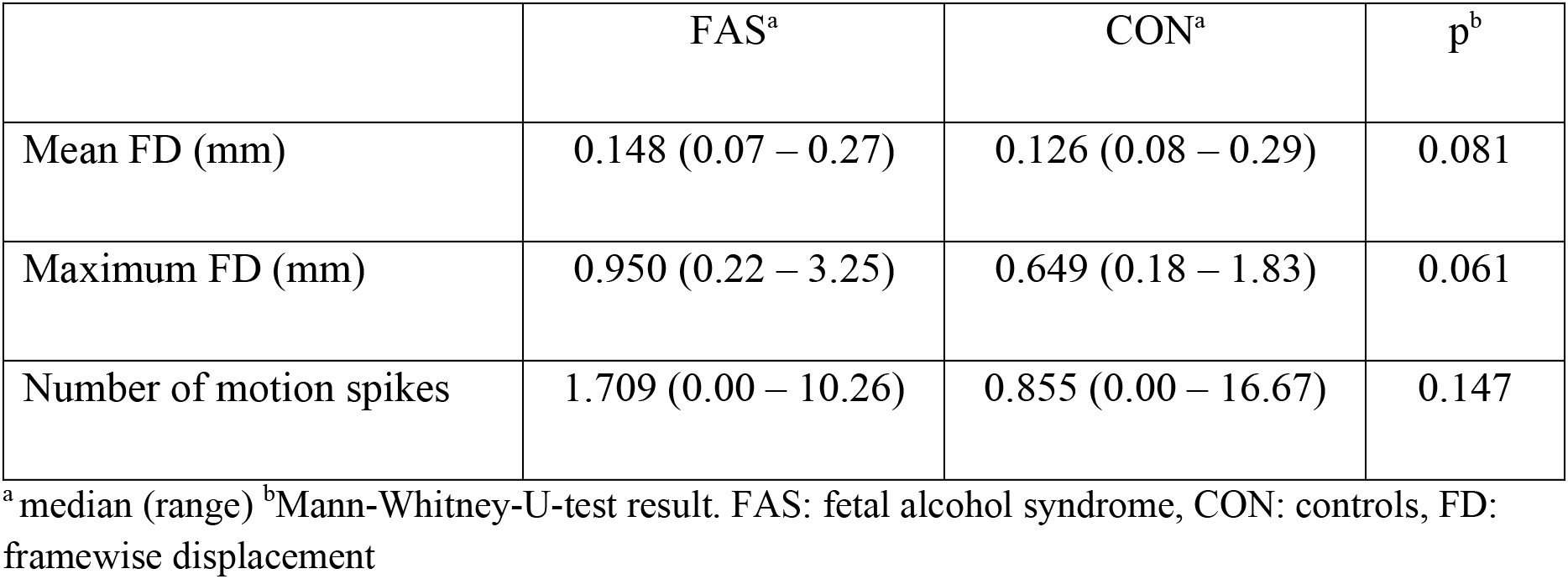
Group comparison of head motion estimates. Data derived from preprocessing (final sample after exclusion of subjects with excessive head motion).

## Supplementary figures

**Supplementary Figure 1.**
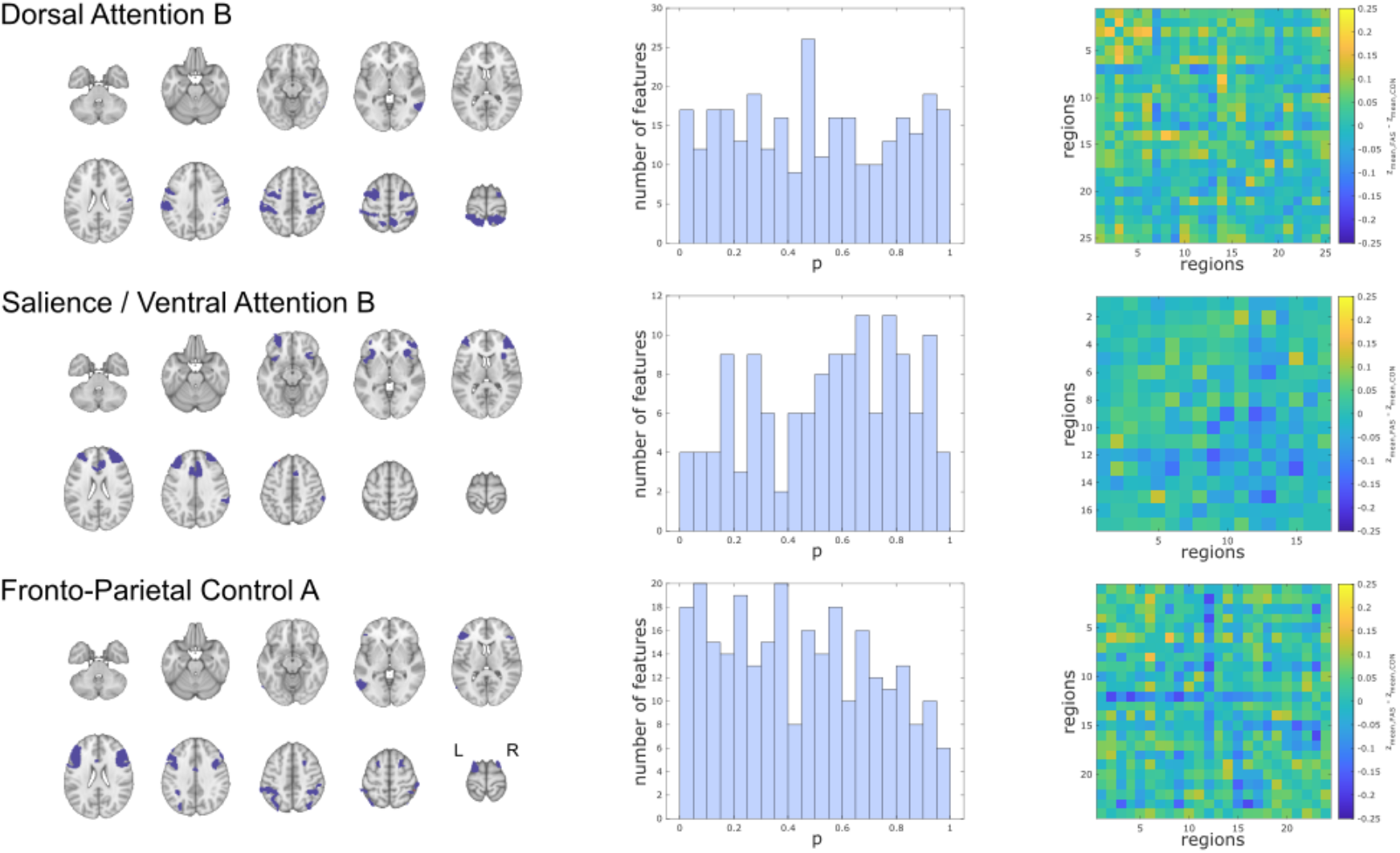
Within-network static functional connectivity of cognition-related brain networks (networks without significant group differences). Three (out of ten) sub-networks not exhibiting altered functional connectivity in FAS patients compared with controls (A – C). Left column: Overview of the networks’ overall extent. Middle column: P-value histograms of multiple two-sided t-tests comparing functional connectivity within these sub-networks between FAS and control subjects (global null hypothesis not rejected based on HC test statistic, Dorsal Attention B: 1.38, Salience / Ventral Attention: 2.30, Fronto-Parietal Control A: 2.92). Right column: Unthresholded matrix of connectivity group differences describing the full connections of cognition- related brain regions within each network. Yellow: mean z-transformed correlation coefficients relatively increased in FAS compared with controls. Blue: relatively decreased functional connectivity in FAS.

**Supplementary Figure 2.**
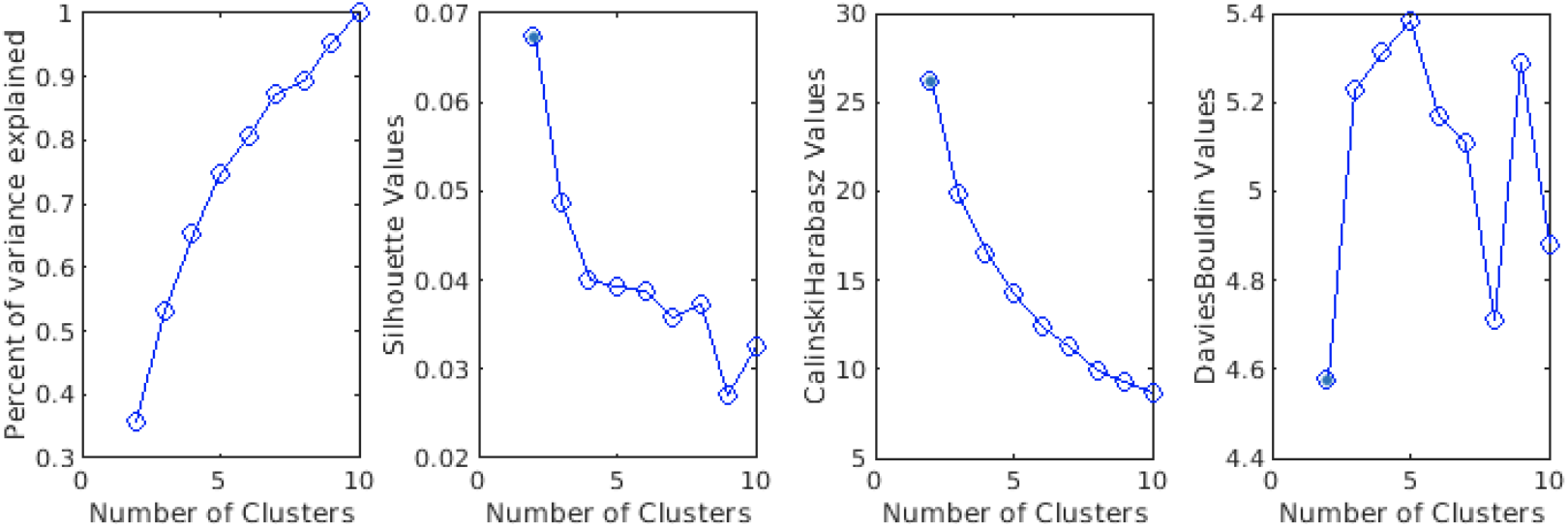
Goodness-of-fit statistics for different k-means clustering solutions (2 to 10 clusters) for putative functional connectivity states in the time-resolved analysis. From left to right: percent of variance explained (relative to a 10-cluster solution), silhouette values (possible range: -1 to 1) with higher values representing good cluster separability, higher Calinski-Harabasz values representing higher cluster density and separability, and Davies–Bouldin criterion (lower values representing higher clustering quality).

**Supplementary Figure 3.**
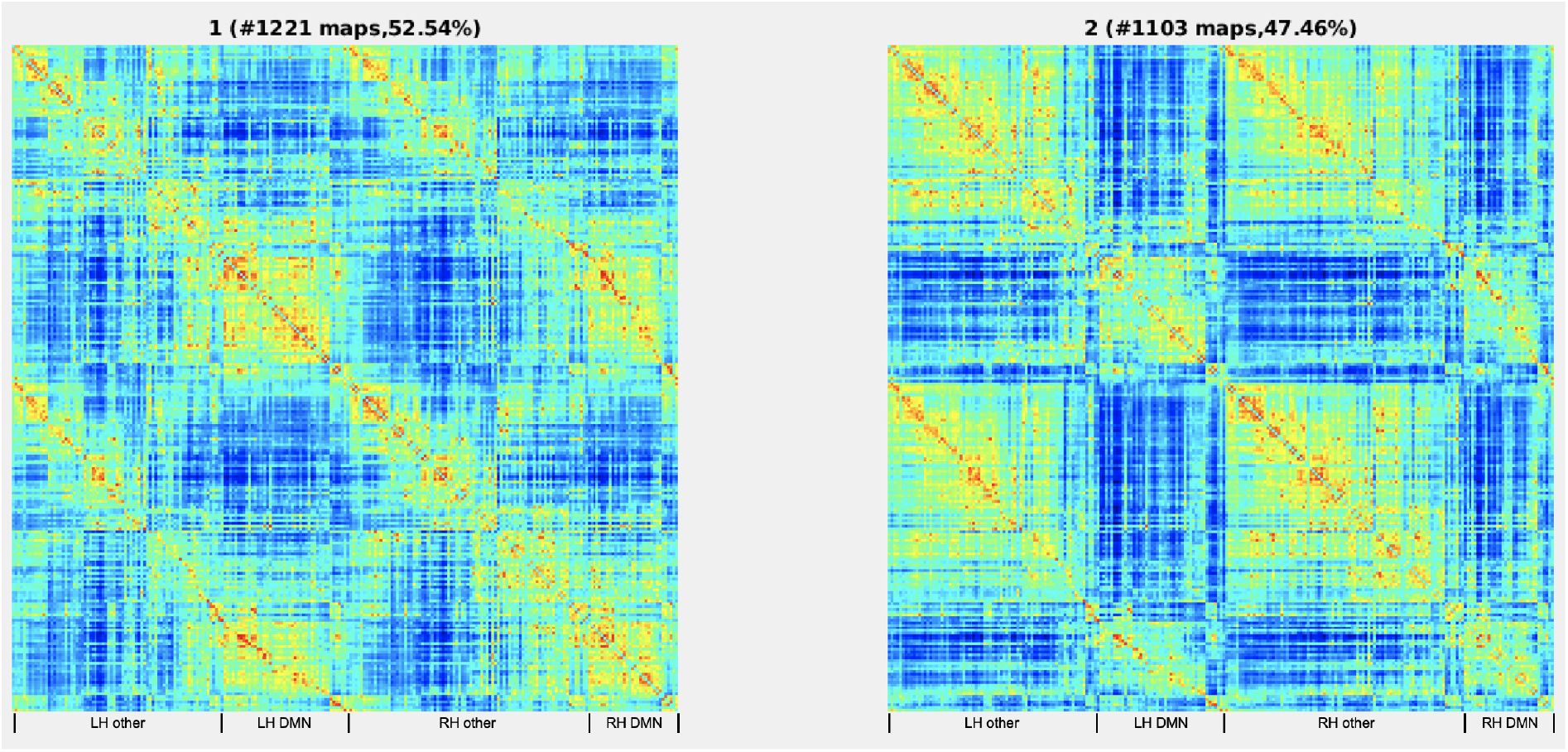
Connectivity matrices representing the two putative functional connectivity (FC) states in the optimal clustering solution of the time-resolved analysis across the entire sample. Original model estimates from the DynamicBC toolbox calculated in the entire sample. Matrices ordered according to the ROI order key provided in the Supplementary data. Left: Cluster 1 shows widely distributed FC dominated by the default mode network (DMN); Right: Cluster 2 exhibits stronger dichotomisation between the DMN and the other cognition-related networks. LH: left hemisphere, RH: right hemisphere

## References

1. Davis KM, Gagnier KR, Moore TE, Todorow M. Cognitive aspects of fetal alcohol spectrum disorder. Wiley Interdiscip Rev Cogn Sci. Jan 2013;4(1):81–92. doi:10.1002/wcs.1202

2. Popova S, Charness ME, Burd L, et al. Fetal alcohol spectrum disorders. Nat Rev Dis Primers. Feb 23 2023;9(1):11. doi:10.1038/s41572-023-00420-x

3. Kodituwakku P, Coriale G, Fiorentino D, et al. Neurobehavioral characteristics of children with fetal alcohol spectrum disorders in communities from Italy: Preliminary results. Alcohol Clin Exp Res. Sep 2006;30(9):1551–61. doi:10.1111/j.1530-0277.2006.00187.x

4. Dorrie N, Focker M, Freunscht I, Hebebrand J. Fetal alcohol spectrum disorders. Eur Child Adolesc Psychiatry. Oct 2014;23(10):863–75. doi:10.1007/s00787-014-0571-6

5. Day NL, Helsel A, Sonon K, Goldschmidt L. The association between prenatal alcohol exposure and behavior at 22 years of age. Alcohol Clin Exp Res. Jul 2013;37(7):1171–8. doi:10.1111/acer.12073

6. Moore EM, Riley EP. What Happens When Children with Fetal Alcohol Spectrum Disorders Become Adults? Curr Dev Disord Rep. Sep 2015;2(3):219–227. doi:10.1007/s40474-015-0053-7

7. Wozniak JR, Riley EP, Charness ME. Clinical presentation, diagnosis, and management of fetal alcohol spectrum disorder. Lancet Neurol. Aug 2019;18(8):760–770. doi:10.1016/S1474-4422(19)30150-4

8. Hoyme HE, Kalberg WO, Elliott AJ, et al. Updated Clinical Guidelines for Diagnosing Fetal Alcohol Spectrum Disorders. Pediatrics. Aug 2016;138(2)doi:10.1542/peds.2015-4256

9. Donald KA, Eastman E, Howells FM, et al. Neuroimaging effects of prenatal alcohol exposure on the developing human brain: a magnetic resonance imaging review. Acta Neuropsychiatr. Oct 2015;27(5):251–69. doi:10.1017/neu.2015.12

10. Niendam TA, Laird AR, Ray KL, Dean YM, Glahn DC, Carter CS. Meta-analytic evidence for a superordinate cognitive control network subserving diverse executive functions. Cogn Affect Behav Neurosci. Jun 2012;12(2):241–68. doi:10.3758/s13415-011-0083-5

11. Menon V. Large-scale brain networks and psychopathology: a unifying triple network model. Trends Cogn Sci. Oct 2011;15(10):483–506. doi:10.1016/j.tics.2011.08.003

12. Duncan J. The multiple-demand (MD) system of the primate brain: mental programs for intelligent behaviour. Trends Cogn Sci. Apr 2010;14(4):172–9. doi:10.1016/j.tics.2010.01.004

13. Smith SM, Fox PT, Miller KL, et al. Correspondence of the brain’s functional architecture during activation and rest. Proc Natl Acad Sci U S A. Aug 4 2009;106(31):13040–5. doi:10.1073/pnas.0905267106

14. van den Heuvel MP, Hulshoff Pol HE. Exploring the brain network: a review on resting-state fMRI functional connectivity. Eur Neuropsychopharmacol. Aug 2010;20(8):519–34. doi:10.1016/j.euroneuro.2010.03.008

15. Iraji A, Faghiri A, Lewis N, Fu Z, Rachakonda S, Calhoun VD. Tools of the trade: estimating time-varying connectivity patterns from fMRI data. Soc Cogn Affect Neurosci. Aug 5 2021;16(8):849–874. doi:10.1093/scan/nsaa114

16. Vidaurre D, Smith SM, Woolrich MW. Brain network dynamics are hierarchically organized in time. Proc Natl Acad Sci U S A. Nov 28 2017;114(48):12827–12832. doi:10.1073/pnas.1705120114

17. Preti MG, Bolton TA, Van De Ville D. The dynamic functional connectome: State-of-the-art and perspectives. Neuroimage. Oct 15 2017;160:41–54. doi:10.1016/j.neuroimage.2016.12.061

18. Fong AHC, Yoo K, Rosenberg MD, et al. Dynamic functional connectivity during task performance and rest predicts individual differences in attention across studies. Neuroimage. Mar 2019;188:14–25. doi:10.1016/j.neuroimage.2018.11.057

19. Madhyastha TM, Askren MK, Boord P, Grabowski TJ. Dynamic connectivity at rest predicts attention task performance. Brain Connect. Feb 2015;5(1):45–59. doi:10.1089/brain.2014.0248

20. Abdallah M, Farrugia N, Chirokoff V, Chanraud S. Static and dynamic aspects of cerebro-cerebellar functional connectivity are associated with self-reported measures of impulsivity: A resting-state fMRI study. Netw Neurosci. 2020;4(3):891–909. doi:10.1162/netn_a_00149

21. Rafi H, Delavari F, Perroud N, Derome M, Debbane M. The continuum of attention dysfunction: Evidence from dynamic functional network connectivity analysis in neurotypical adolescents. PLoS One. 2023;18(1):e0279260. doi:10.1371/journal.pone.0279260

22. Yang Y, Yang B, Zhang L, Peng G, Fang D. Dynamic Functional Connectivity Reveals Abnormal Variability in the Amygdala Subregions of Children With Attention-Deficit/Hyperactivity Disorder. Front Neurosci. 2021;15:648143. doi:10.3389/fnins.2021.648143

23. Ahmadi M, Kazemi K, Kuc K, Cybulska-Klosowicz A, Helfroush MS, Aarabi A. Resting state dynamic functional connectivity in children with attention deficit/hyperactivity disorder. J Neural Eng. Aug 16 2021;18(4) doi:10.1088/1741-2552/ac16b3

24. Agoalikum E, Klugah-Brown B, Yang H, et al. Differences in Disrupted Dynamic Functional Network Connectivity Among Children, Adolescents, and Adults With Attention Deficit/Hyperactivity Disorder: A Resting-State fMRI Study. Front Hum Neurosci. 2021;15:697696. doi:10.3389/fnhum.2021.697696

25. Fan J, Taylor PA, Jacobson SW, et al. Localized reductions in resting-state functional connectivity in children with prenatal alcohol exposure. Hum Brain Mapp. Oct 2017;38(10):5217–5233. doi:10.1002/hbm.23726

26. Little G, Reynolds J, Beaulieu C. Altered Functional Connectivity Observed at Rest in Children and Adolescents Prenatally Exposed to Alcohol. Brain Connect. Oct 2018;8(8):503–515. doi:10.1089/brain.2017.0572

27. Ware AL, Long X, Lebel C. Functional connectivity of the attention networks is altered and relates to neuropsychological outcomes in children with prenatal alcohol exposure. Dev Cogn Neurosci. Apr 2021;48:100951. doi:10.1016/j.dcn.2021.100951

28. Wozniak JR, Mueller BA, Mattson SN, et al. Functional connectivity abnormalities and associated cognitive deficits in fetal alcohol Spectrum disorders (FASD). Brain Imaging Behav. Oct 2017;11(5):1432–1445. doi:10.1007/s11682-016-9624-4

29. Wozniak JR, Mueller BA, Bell CJ, et al. Global functional connectivity abnormalities in children with fetal alcohol spectrum disorders. Alcohol Clin Exp Res. May 2013;37(5):748–56. doi:10.1111/acer.12024

30. Long X, Kar P, Gibbard B, Tortorelli C, Lebel C. The brain’s functional connectome in young children with prenatal alcohol exposure. Neuroimage Clin. 2019;24:102082. doi:10.1016/j.nicl.2019.102082

31. Rodriguez CI, Vergara VM, Calhoun VD, et al. Disruptions in global network segregation and integration in adolescents and young adults with fetal alcohol spectrum disorder. Alcohol Clin Exp Res. Sep 2021;45(9):1775–1789. doi:10.1111/acer.14673

32. Santhanam P, Coles CD, Li Z, Li L, Lynch ME, Hu X. Default mode network dysfunction in adults with prenatal alcohol exposure. Psychiatry Res. Dec 30 2011;194(3):354–362. doi:10.1016/j.pscychresns.2011.05.004

33. Wozniak JR, Mueller BA, Muetzel RL, et al. Inter-hemispheric functional connectivity disruption in children with prenatal alcohol exposure. Alcohol Clin Exp Res. May 2011;35(5):849–61. doi:10.1111/j.1530-0277.2010.01415.x

34. Long X, Little G, Beaulieu C, Lebel C. Sensorimotor network alterations in children and youth with prenatal alcohol exposure. Hum Brain Mapp. May 2018;39(5):2258–2268. doi:10.1002/hbm.24004

35. Sundermann B, Garde S, Dehghan Nayyeri M, et al. Approaching altered inhibitory control in phenylketonuria: A functional MRI study with a Go-NoGo task in young female adults. Eur J Neurosci. Oct 2020;52(8):3951–3962. doi:10.1111/ejn.14738

36. Rau JMH, Sundermann B, Pfleiderer B, et al. Inhibitory control in young adult women with fetal alcohol syndrome: Findings from a pilot functional magnetic resonance imaging study. Alcohol: Clinical and Experimental Research. 2023/02/17 2023;n/a doi:10.1111/acer.15025

37. Majewski F. Untersuchungen zur Alkoholembryopathie. Thieme Stuttgart/New York; 1980.

38. Famy C, Streissguth AP, Unis AS. Mental illness in adults with fetal alcohol syndrome or fetal alcohol effects. Am J Psychiatry. Apr 1998;155(4):552–4. doi:10.1176/ajp.155.4.552

39. Sundermann B, Pfleiderer B, Minnerup H, Berger K, Douaud G. Interaction of Developmental Venous Anomalies with Resting-State Functional MRI Measures. AJNR Am J Neuroradiol. Dec 2018;39(12):2326–2331. doi:10.3174/ajnr.A5847

40. Dosenbach NU, Nardos B, Cohen AL, et al. Prediction of individual brain maturity using fMRI. Science. Sep 10 2010;329(5997):1358–61. doi:10.1126/science.1194144

41. Oldfield RC. The assessment and analysis of handedness: the Edinburgh inventory. Neuropsychologia. Mar 1971;9(1):97–113. doi:10.1016/0028-3932(71)90067-4

42. Oswald WD, Roth E. Der Zahlenverbindungstest (ZVT). Ein sprachfreier Intelligenz-Test zur Messung der “kognitiven Leistungsgeschwindigkeit*”*. . 2nd ed. Hogrefe; 1987.

43. Wittchen HU, Pfister H. DIA-X-Interviews: Manual für Screening-Verfahren und Interview; Interviewheft Längsschnittuntersuchung (DIA-X-Lifetime); Ergänzungsheft (DIA-X-Lifetime); Interviewheft Querschnittuntersuchung (DIA-X-12 Monate); Ergänzungsheft (DIA-X-12 Monate); PC-Programm zur Durchführung des Interviews (Längs- und Querschnittuntersuchung); Auswertungsprogramm. Swets & Zeitlinger; 1997.

44. Salthouse TA. What cognitive abilities are involved in trail-making performance? Intelligence. Jul 2011;39(4):222–232. doi:10.1016/j.intell.2011.03.001

45. Gorgolewski KJ, Auer T, Calhoun VD, et al. The brain imaging data structure, a format for organizing and describing outputs of neuroimaging experiments. Sci Data. Jun 21 2016;3:160044. doi:10.1038/sdata.2016.44

46. Wulms N. BiDirect-BIDS-ConverteR. 2019;doi:10.5281/zenodo.3469538

47. Bischoff-Grethe A, Ozyurt IB, Busa E, et al. A technique for the deidentification of structural brain MR images. Hum Brain Mapp. Sep 2007;28(9):892–903. doi:10.1002/hbm.20312

48. Esteban O, Markiewicz CJ, Blair RW, et al. fMRIPrep: a robust preprocessing pipeline for functional MRI. Nat Methods. Jan 2019;16(1):111–116. doi:10.1038/s41592-018-0235-4

49. Finc KC, M.; Bonna, K. fMRIDenoise: automated denoising, denoising strategies comparison, and functional connectivity data quality control. 2021;doi:10.5281/zenodo.4458072

50. Satterthwaite TD, Elliott MA, Gerraty RT, et al. An improved framework for confound regression and filtering for control of motion artifact in the preprocessing of resting-state functional connectivity data. Neuroimage. Jan 1 2013;64:240–56. doi:10.1016/j.neuroimage.2012.08.052

51. Power JD, Barnes KA, Snyder AZ, Schlaggar BL, Petersen SE. Spurious but systematic correlations in functional connectivity MRI networks arise from subject motion. Neuroimage. Feb 1 2012;59(3):2142–54. doi:10.1016/j.neuroimage.2011.10.018

52. Ciric R, Thompson WH, Lorenz R, et al. TemplateFlow: FAIR-sharing of multi-scale, multi-species brain models. Nat Methods. Dec 2022;19(12):1568–1571. doi:10.1038/s41592-022-01681-2

53. Schaefer A, Kong R, Gordon EM, et al. Local-Global Parcellation of the Human Cerebral Cortex from Intrinsic Functional Connectivity MRI. Cereb Cortex. Sep 1 2018;28(9):3095–3114. doi:10.1093/cercor/bhx179

54. Yeo BT, Krienen FM, Sepulcre J, et al. The organization of the human cerebral cortex estimated by intrinsic functional connectivity. J Neurophysiol. Sep 2011;106(3):1125–65. doi:10.1152/jn.00338.2011

55. Chao-Gan Y, Yu-Feng Z. DPARSF: A MATLAB Toolbox for “Pipeline” Data Analysis of Resting-State fMRI. Front Syst Neurosci. 2010;4:13. doi:10.3389/fnsys.2010.00013

56. Donoho D, Jin J. Higher Criticism for Large-Scale Inference, Especially for Rare and Weak Effects. Statistical Science. 2/1 2015;30(1):1-25. doi:10.1214/14-STS506

57. Donoho D, Jin J. Higher criticism for detecting sparse heterogeneous mixtures. The Annals of Statistics. 6/1 2004;32(3):962-994. doi:10.1214/009053604000000265

58. Breheny P, Stromberg A, Lambert J. p-Value Histograms: Inference and Diagnostics. High Throughput. Aug 31 2018;7(3)doi:10.3390/ht7030023

59. Cai TT, Sun W. Large-Scale Global and Simultaneous Inference: Estimation and Testing in Very High Dimensions. Annual Review of Economics. 2017/08/02 2017;9(1):411–439. doi:10.1146/annurev-economics-063016-104355

60. Gerlach AR, Karim HT, Kazan J, Aizenstein HJ, Krafty RT, Andreescu C. Networks of worry-towards a connectivity-based signature of late-life worry using higher criticism. Transl Psychiatry. Oct 28 2021;11(1):550. doi:10.1038/s41398-021-01648-5

61. Yekutieli D, Benjamini Y. Resampling-based false discovery rate controlling multiple test procedures for correlated test statistics. Journal of Statistical Planning and Inference. 1999/12/01/ 1999;82(1):171–196. doi:10.1016/S0378-3758(99)00041-5

62. Liao W, Wu GR, Xu Q, et al. DynamicBC: a MATLAB toolbox for dynamic brain connectome analysis. Brain Connect. Dec 2014;4(10):780–90. doi:10.1089/brain.2014.0253

63. Damaraju E, Allen EA, Belger A, et al. Dynamic functional connectivity analysis reveals transient states of dysconnectivity in schizophrenia. Neuroimage Clin. 2014;5:298–308. doi:10.1016/j.nicl.2014.07.003

64. Li J, Zhang D, Liang A, et al. High transition frequencies of dynamic functional connectivity states in the creative brain. Sci Rep. Apr 6 2017;7:46072. doi:10.1038/srep46072

65. Caliński T, Harabasz J. A dendrite method for cluster analysis. Communications in Statistics. 1974/01/01 1974;3(1):1-27. doi:10.1080/03610927408827101

66. Davies DL, Bouldin DW. A Cluster Separation Measure. IEEE Transactions on Pattern Analysis and Machine Intelligence. 1979;PAMI-1(2):224–227. doi:10.1109/TPAMI.1979.4766909

67. Rousseeuw PJ. Silhouettes: A graphical aid to the interpretation and validation of cluster analysis. Journal of Computational and Applied Mathematics. 1987/11/01/ 1987;20:53–65. doi:10.1016/0377-0427(87)90125-7

68. Coles CD, Platzman KA, Lynch ME, Freides D. Auditory and visual sustained attention in adolescents prenatally exposed to alcohol. Alcohol Clin Exp Res. Feb 2002;26(2):263–71.

69. Gautam P, Nunez SC, Narr KL, et al. Developmental Trajectories for Visuo-Spatial Attention are Altered by Prenatal Alcohol Exposure: A Longitudinal FMRI Study. Cereb Cortex. Dec 2015;25(12):4761–71. doi:10.1093/cercor/bhu162

70. Kooistra L, Crawford S, Gibbard B, Ramage B, Kaplan BJ. Differentiating attention deficits in children with fetal alcohol spectrum disorder or attention-deficit-hyperactivity disorder. Dev Med Child Neurol. Feb 2010;52(2):205–11. doi:10.1111/j.1469-8749.2009.03352.x

71. Brown RT, Coles CD, Smith IE, et al. Effects of prenatal alcohol exposure at school age. II. Attention and behavior. Neurotoxicol Teratol. Jul-Aug 1991;13(4):369–76. doi:10.1016/0892-0362(91)90085-b

72. Infante MA, Moore EM, Nguyen TT, Fourligas N, Mattson SN, Riley EP. Objective assessment of ADHD core symptoms in children with heavy prenatal alcohol exposure. Physiol Behav. Sep 1 2015;148:45–50. doi:10.1016/j.physbeh.2014.10.014

73. Connor PD, Streissguth AP, Sampson PD, Bookstein FL, Barr HM. Individual differences in auditory and visual attention among fetal alcohol-affected adults. Alcohol Clin Exp Res. Aug 1999;23(8):1395–402.

74. Hutzler F. Reverse inference is not a fallacy per se: cognitive processes can be inferred from functional imaging data. Neuroimage. Jan 1 2014;84:1061–9. doi:10.1016/j.neuroimage.2012.12.075

75. Poldrack RA. Can cognitive processes be inferred from neuroimaging data? Trends Cogn Sci. Feb 2006;10(2):59–63. doi:10.1016/j.tics.2005.12.004

76. Candelaria-Cook FT, Schendel ME, Flynn L, Hill DE, Stephen JM. Altered Resting-State Neural Oscillations and Spectral Power in Children with Fetal Alcohol Spectrum Disorder. Alcohol Clin Exp Res. Jan 2021;45(1):117–130. doi:10.1111/acer.14502

77. Long X, Little G, Treit S, Beaulieu C, Gong G, Lebel C. Altered brain white matter connectome in children and adolescents with prenatal alcohol exposure. Brain Struct Funct. Apr 2020;225(3):1123–1133. doi:10.1007/s00429-020-02064-z

78. Zanto TP, Gazzaley A. Fronto-parietal network: flexible hub of cognitive control. Trends Cogn Sci. Dec 2013;17(12):602–3. doi:10.1016/j.tics.2013.10.001

79. Kodali VN, Jacobson JL, Lindinger NM, et al. Differential Recruitment of Brain Regions During Response Inhibition in Children Prenatally Exposed to Alcohol. Alcohol Clin Exp Res. Feb 2017;41(2):334–344. doi:10.1111/acer.13307

80. Infante MA, Moore EM, Bischoff-Grethe A, Tapert SF, Mattson SN, Riley EP. Altered functional connectivity during spatial working memory in children with heavy prenatal alcohol exposure. Alcohol. Nov 2017;64:11–21. doi:10.1016/j.alcohol.2017.05.002

81. Fox MD, Snyder AZ, Vincent JL, Corbetta M, Van Essen DC, Raichle ME. The human brain is intrinsically organized into dynamic, anticorrelated functional networks. Proc Natl Acad Sci U S A. Jul 5 2005;102(27):9673–8. doi:10.1073/pnas.0504136102

82. Smith V, Mitchell DJ, Duncan J. Role of the Default Mode Network in Cognitive Transitions. Cereb Cortex. Oct 1 2018;28(10):3685–3696. doi:10.1093/cercor/bhy167

83. Crittenden BM, Mitchell DJ, Duncan J. Recruitment of the default mode network during a demanding act of executive control. Elife. Apr 13 2015;4:e06481. doi:10.7554/eLife.06481

84. Andrews-Hanna JR, Reidler JS, Sepulcre J, Poulin R, Buckner RL. Functional-anatomic fractionation of the brain’s default network. Neuron. Feb 25 2010;65(4):550–62. doi:10.1016/j.neuron.2010.02.005

85. Popova S, Lange S, Probst C, Gmel G, Rehm J. Estimation of national, regional, and global prevalence of alcohol use during pregnancy and fetal alcohol syndrome: a systematic review and meta-analysis. Lancet Glob Health. Mar 2017;5(3):e290–e299. doi:10.1016/S2214-109X(17)30021-9

86. Cremers HR, Wager TD, Yarkoni T. The relation between statistical power and inference in fMRI. PLoS One. 2017;12(11):e0184923. doi:10.1371/journal.pone.0184923

87. Ioannidis JP. Why most published research findings are false. PLoS Med. Aug 2005;2(8):e124. doi:10.1371/journal.pmed.0020124

88. Yarkoni T. Big Correlations in Little Studies: Inflated fMRI Correlations Reflect Low Statistical Power- Commentary on Vul et al. (2009). Perspect Psychol Sci. May 2009;4(3):294–8. doi:10.1111/j.1745-6924.2009.01127.x

89. Yarkoni T, Braver TS. Cognitive Neuroscience Approaches to Individual Differences in Working Memory and Executive Control: Conceptual and Methodological Issues. In: Gruszka A, Matthews G, Szymura B, eds. Handbook of Individual Differences in Cognition: Attention, Memory, and Executive Control. Springer New York; 2010:87–107.

90. Chen G, Taylor PA, Cox RW. Is the statistic value all we should care about in neuroimaging? Neuroimage. Feb 15 2017;147:952–959. doi:10.1016/j.neuroimage.2016.09.066

91. Taylor PA, Reynolds RC, Calhoun V, et al. Highlight Results, Don’t Hide Them: Enhance interpretation, reduce biases and improve reproducibility. NeuroImage. 2023;274:120138. doi:10.1016/j.neuroimage.2023.120138

92. Sundermann B, Pfleiderer B, McLeod A, Mathys C. Seeing more than the tip of the iceberg: Approaches to subthreshold effects in functional magnetic resonance imaging of the brain. PsyArXiv Preprints. May 7 2023;doi:10.31234/osf.io/fyhst

93. Gorgolewski KJ, Varoquaux G, Rivera G, et al. NeuroVault.org: a web-based repository for collecting and sharing unthresholded statistical maps of the human brain. Front Neuroinform. 2015;9:8. doi:10.3389/fninf.2015.00008

94. Glass L, Graham DM, Deweese BN, Jones KL, Riley EP, Mattson SN. Correspondence of parent report and laboratory measures of inattention and hyperactivity in children with heavy prenatal alcohol exposure. Neurotoxicol Teratol. Mar-Apr 2014;42:43–50. doi:10.1016/j.ntt.2014.01.007

95. Long X, Lebel C. Evaluation of Brain Alterations and Behavior in Children With Low Levels of Prenatal Alcohol Exposure. JAMA Netw Open. Apr 1 2022;5(4):e225972. doi:10.1001/jamanetworkopen.2022.5972

96. Lees B, Mewton L, Jacobus J, et al. Association of Prenatal Alcohol Exposure With Psychological, Behavioral, and Neurodevelopmental Outcomes in Children From the Adolescent Brain Cognitive Development Study. Am J Psychiatry. Nov 1 2020;177(11):1060–1072. doi:10.1176/appi.ajp.2020.20010086

97. Craddock RC, James GA, Holtzheimer PE, 3rd, Hu XP, Mayberg HS. A whole brain fMRI atlas generated via spatially constrained spectral clustering. Hum Brain Mapp. Aug 2012;33(8):1914–28. doi:10.1002/hbm.21333

98. Alves PN, Foulon C, Karolis V, et al. An improved neuroanatomical model of the default-mode network reconciles previous neuroimaging and neuropathological findings. Commun Biol. 2019;2:370. doi:10.1038/s42003-019-0611-3

99. Sundermann B, Pfleiderer B. Functional connectivity profile of the human inferior frontal junction: involvement in a cognitive control network. BMC Neurosci. Oct 3 2012;13:119. doi:10.1186/1471-2202-13-119

100. Uddin LQ, Yeo BTT, Spreng RN. Towards a Universal Taxonomy of Macro-scale Functional Human Brain Networks. Brain Topogr. Nov 2019;32(6):926–942. doi:10.1007/s10548-019-00744-6

101. Li J, Siegmund D. Higher criticism: p-values and criticism. The Annals of Statistics. 2015;43(3):1323–1350, 28. doi:10.1214/15-AOS1312

102. Carp J. On the plurality of (methodological) worlds: estimating the analytic flexibility of FMRI experiments. Front Neurosci. 2012;6:149. doi:10.3389/fnins.2012.00149

103. Hindriks R, Adhikari MH, Murayama Y, et al. Can sliding-window correlations reveal dynamic functional connectivity in resting-state fMRI? Neuroimage. Feb 15 2016;127:242–256. doi:10.1016/j.neuroimage.2015.11.055

104. Allen EA, Damaraju E, Plis SM, Erhardt EB, Eichele T, Calhoun VD. Tracking whole-brain connectivity dynamics in the resting state. Cereb Cortex. Mar 2014;24(3):663–76. doi:10.1093/cercor/bhs352

## References

Abraham, Alexandre, Fabian Pedregosa, Michael Eickenberg, Philippe Gervais, Andreas Mueller, Jean Kossaifi, Alexandre Gramfort, Bertrand Thirion, and Gael Varoquaux. 2014. “Machine Learning for Neuroimaging with Scikit-Learn.” Frontiers in Neuroinformatics 8. https://doi.org/10.3389/fninf.2014.00014.

Avants, B.B., C.L. Epstein, M. Grossman, and J.C. Gee. 2008. “Symmetric Diffeomorphic Image Registration with Cross-Correlation: Evaluating Automated Labeling of Elderly and Neurodegenerative Brain.” Medical Image Analysis 12 (1): 26–41. https://doi.org/10.1016/j.media.2007.06.004.

Behzadi, Yashar, Khaled Restom, Joy Liau, and Thomas T. Liu. 2007. “A Component Based Noise Correction Method (CompCor) for BOLD and Perfusion Based fMRI.” NeuroImage 37 (1): 90–101. https://doi.org/10.1016/j.neuroimage.2007.04.042.

Dale, Anders M., Bruce Fischl, and Martin I. Sereno. 1999. “Cortical Surface-Based Analysis: I. Segmentation and Surface Reconstruction.” NeuroImage 9 (2): 179–94. https://doi.org/10.1006/nimg.1998.0395.

Esteban, Oscar, Ross Blair, Christopher J. Markiewicz, Shoshana L. Berleant, Craig Moodie, Feilong Ma, Ayse Ilkay Isik, et al. 2018. “FMRIPrep.” Software. Zenodo. https://doi.org/10.5281/zenodo.852659.

Esteban, Oscar, Christopher Markiewicz, Ross W Blair, Craig Moodie, Ayse Ilkay Isik, Asier Erramuzpe Aliaga, James Kent, et al. 2018. “fMRIPrep: A Robust Preprocessing Pipeline for Functional MRI.” Nature Methods. https://doi.org/10.1038/s41592-018-0235-4.

Fonov, VS, AC Evans, RC McKinstry, CR Almli, and DL Collins. 2009. “Unbiased Nonlinear Average Age-Appropriate Brain Templates from Birth to Adulthood.” NeuroImage 47, Supplement 1: S102. https://doi.org/10.1016/S1053-8119(09)70884-5.

Gorgolewski, K., C. D. Burns, C. Madison, D. Clark, Y. O. Halchenko, M. L. Waskom, and S. Ghosh. 2011. “Nipype: A Flexible, Lightweight and Extensible Neuroimaging Data Processing Framework in Python.” Frontiers in Neuroinformatics 5: 13. https://doi.org/10.3389/fninf.2011.00013.

Gorgolewski, Krzysztof J., Oscar Esteban, Christopher J. Markiewicz, Erik Ziegler, David Gage Ellis, Michael Philipp Notter, Dorota Jarecka, et al. 2018. “Nipype.” Software. Zenodo. https://doi.org/10.5281/zenodo.596855.

Greve, Douglas N, and Bruce Fischl. 2009. “Accurate and Robust Brain Image Alignment Using Boundary-Based Registration.” NeuroImage 48 (1): 63–72. https://doi.org/10.1016/j.neuroimage.2009.06.060.

Jenkinson, Mark, Peter Bannister, Michael Brady, and Stephen Smith. 2002. “Improved Optimization for the Robust and Accurate Linear Registration and Motion Correction of Brain Images.” NeuroImage 17 (2): 825–41. https://doi.org/10.1006/nimg.2002.1132.

Klein, Arno, Satrajit S. Ghosh, Forrest S. Bao, Joachim Giard, Yrjö Häme, Eliezer Stavsky, Noah Lee, et al. 2017. “Mindboggling Morphometry of Human Brains.” PLOS Computational Biology 13 (2): e1005350. https://doi.org/10.1371/journal.pcbi.1005350.

Lanczos, C. 1964. “Evaluation of Noisy Data.” Journal of the Society for Industrial and Applied Mathematics Series B Numerical Analysis 1 (1): 76–85. https://doi.org/10.1137/0701007.

Power, Jonathan D., Anish Mitra, Timothy O. Laumann, Abraham Z. Snyder, Bradley L. Schlaggar, and Steven E. Petersen. 2014. “Methods to Detect, Characterize, and Remove Motion Artifact in Resting State fMRI.” NeuroImage 84 (Supplement C): 320–41. https://doi.org/10.1016/j.neuroimage.2013.08.048.

Satterthwaite, Theodore D., Mark A. Elliott, Raphael T. Gerraty, Kosha Ruparel, James Loughead, Monica E. Calkins, Simon B. Eickhoff, et al. 2013. “An improved framework for confound regression and filtering for control of motion artifact in the preprocessing of resting-state functional connectivity data.” NeuroImage 64 (1): 240–56. https://doi.org/10.1016/j.neuroimage.2012.08.052.

Tustison, N. J., B. B. Avants, P. A. Cook, Y. Zheng, A. Egan, P. A. Yushkevich, and J. C. Gee. 2010. “N4ITK: Improved N3 Bias Correction.” IEEE Transactions on Medical Imaging 29 (6): 1310–20. https://doi.org/10.1109/TMI.2010.2046908.

Zhang, Y., M. Brady, and S. Smith. 2001. “Segmentation of Brain MR Images Through a Hidden Markov Random Field Model and the Expectation-Maximization Algorithm.” IEEE Transactions on Medical Imaging 20 (1): 45–57. https://doi.org/10.1109/42.906424.

